# Identifying autism in women diagnosed with borderline personality disorder: Clinician and lived experience perspectives

**DOI:** 10.64898/2026.04.27.26351287

**Authors:** J. Parker, E Thompson, W Mandy, R McCabe, E. Stark, K Barnicot

**Affiliations:** Department of Population Health and Policy, City St George’s, University of London. United Kingdom; Department of Clinical, Educational and Health Psychology, University College London. United Kingdom; Oxford Health NHS Foundation Trust, Warneford Hospital, Oxford. United Kingdom; Centre for Eudaimonia and Human Flourishing, Linacre College, University of Oxford, Oxford. United Kingdom

**Keywords:** Autism, personality disorder, diagnosis, misdiagnosis, lived experience, qualitative

## Abstract

**Background:** Growing numbers of people with a borderline personality disorder (BPD) diagnosis are realising they may have undiagnosed autism. Previous qualitative research has not focused on identifying barriers and facilitators to this diagnostic journey, did not explore the perspectives of clinicians, and did not include the experiences of people who are unsure whether they are autistic or not. We aimed to understand lived experience and clinician perspectives on facilitators and barriers to recognising undiagnosed autism, in women and people assigned female at birth (PAFAB) with a diagnosis of personality disorder.

**Methods:** We carried out in-depth qualitative interviews with 15 mental health clinicians, and 15 women/PAFAB who had a current or prior diagnosis of BPD and identified as definitely or possibly autistic, from across the United Kingdom. We analysed the interview data using reflexive thematic analysis.

**Results:** Both clinician and lived experience participants identified many barriers to recognising autism in women and PAFAB with a BPD diagnosis: BPD diagnoses being made with minimal assessment during mental health crises, systemic incentivisation to diagnose BPD in order to access psychological therapies, siloed service pathways, clinician reluctance to question pre-existing BPD diagnoses, pathologizing of patients for questioning their BPD diagnosis, and lack of clinician knowledge about different presentations of autism or about ways that autism presents similarly and differently to BPD. Participants identified numerous ways in which autistic characteristics could be misattributed as symptomatic of BPD, further contributing to missed or misdiagnosis.

**Conclusion:** Our findings suggest that improving clinician awareness of different presentations of autism, and of differential diagnosis from BPD is likely to reduce misdiagnosis, alongside avoiding rapid diagnostic decisions during mental health crises. Our study further highlights the value of being open to questioning pre-existing diagnoses, joint working across autism and personality disorder services, and improving transdiagnostic access to psychological interventions.

**Community Brief:** *Why is this an important issue?:* Autism in adults may be missed, or mis-diagnosed as a mental health condition. Borderline personality disorder (BPD) is the most common perceived misdiagnosis held by autistic people. Unrecognised autism may lead to worsened mental health in BPD-diagnosed people.

*What was the purpose of this study?:* We aimed to understand lived experience and clinician perspectives on what gets in the way of recognising undiagnosed autism, in people with a diagnosis of BPD.

*What did the researchers do?:* We interviewed 15 mental health clinicians, and 15 women/people assigned female at birth (AFAB), who had a current or prior diagnosis of BPD and identified as definitely or possibly autistic. We asked lived experience participants about their experiences of realising that they may be autistic. We asked clinicians to share their experiences of differentiating autism and personality disorder in clinical practice. We asked all participants to discuss their experiences of what makes it challenging to recognise autism in BPD-diagnosed people, and what helps.

*What were the results and conclusions of the study?:* Both clinician and lived experience participants identified many barriers to recognising autism in women and AFAB people with a BPD diagnosis. They said BPD diagnoses are made with minimal assessment during mental health crises. They said clinicians feel that they have to diagnose BPD in order to help people access psychological therapies. They said service pathways separate out autism and BPD rather than considering them together. They said clinicians are reluctant to question pre-existing BPD diagnoses, and that patients’ questioning of their BPD diagnosis is sometimes seen as symptomatic of mental health difficulties. They said clinicians lack knowledge about how autism can look different in women and AFAB people, and about ways that autism can look similar and different to BPD. Participants identified numerous ways in which autistic characteristics could be misattributed as symptomatic of BPD, further contributing to missed or misdiagnosis. We concluded that improving mental health clinicians’ understanding of autism, and how it is different from BPD, may help to improve recognition of autism in BPD-diagnosed people. We also concluded it’s important for clinicians to be open to questioning pre-existing diagnoses, to establish joint working across autism and personality disorder services, and to improve transdiagnostic access to psychological interventions.

*What is new or controversial about these findings?:* It’s the first time clinician and lived experience perspectives on this issue have been brought together. It’s controversial because it suggests that mental health services are sometimes not good at recognising autism in BPD- diagnosed people, and that people are potentially being harmed by this.

*What are potential weaknesses in the study?:* We would have liked to understand more about the experiences of ethnically diverse people. Our study may have attracted people who disagree with the idea of BPD and who believe autism is underdiagnosed.

*How will these findings help autistic adults now or in the future?:* We hope it will help BPD-diagnosed people with undiagnosed autism to be better recognised and understood by mental health services.

## Background

Autism in adults may be missed, or mis-diagnosed as a mental health condition (1, 2). This is particularly likely to occur in autistic women and people assigned female at birth (AFAB), who have a higher number of prior mental health diagnoses than autistic men and are diagnosed later in life (1, 3, 4).

Borderline personality disorder (BPD), defined as “a pervasive pattern of instability of interpersonal relationships, self-image, and affects, and marked impulsivity” (5), is the most common perceived misdiagnosis held by autistic people (6). Autism may be missed in BPD-diagnosed people due to gendered stereotypes, assessment methods for each diagnosis, communication differences whereby autistic people can struggle to communicate with and to be understood by neurotypical clinicians, and similarities in the clinical presentation of each diagnosis (such as autistic individuals presenting with behaviours that appear similar to BPD including emotional dysregulation, and self-harm) (3, 7–9).

Lived experience accounts suggest harm caused by failure to recognise undiagnosed autism, particularly for people given a diagnosis of BPD (10–12). Conversely, autistic people say that identifying undiagnosed autism can lead to increased connectedness, self-compassion and self-understanding (13,14), including amongst people with a prior BPD diagnosis (11, 12).

Two previous qualitative studies have explored the experiences of autistic adults previously diagnosed with BPD (11, 12), but did not focus on identifying barriers and facilitators to this diagnostic journey, did not explore the perspectives of clinicians, and did not include the experiences of people who are unsure whether they are autistic. To address this gap, we aimed to understand lived experience and clinician perspectives on facilitators and barriers to recognising undiagnosed autism, in people with a diagnosis of personality disorder.

## Methods

### Design

Qualitative interviews drawing on a critical realist and reflexive analysis approach (15, 16).

### Ethics statement

The study was ethically reviewed and approved by the West Midlands - South Birmingham Research Ethics Committee (ref. 21/WM/0287) on 14^th^ January 2022. We co-designed all research materials with our lived experience panel. Participants gave written informed consent online, over email, or with a researcher. We mitigated potential participant distress following disclosure of personal experiences by offering post-interview debriefing with the interviewer, and signposting to online and local information and support.

### Eligibility criteria

We included people with lived experience if they were aged 18 to 65 years, identified as female and/or AFAB, had a current or historical personality disorder diagnosis, and identified as definitely or possibly autistic. We excluded people who had a schizophrenia spectrum disorder diagnosis, moderate to severe learning difficulties, and/or lived outside the United Kingdom (UK). We included clinicians if they had worked with autism and/or personality disorder diagnosed individuals in the UK.

### Recruitment and sampling

We recruited lived experience participants between February and December 2022, from autism and personality disorder services in three NHS Trusts in London, North-West and North-Central England, and through social media (‘X’ accounts of the research team and study funders; mailing list held by the research funders). We also invited participation from eligible individuals taking part in a separate study on diagnostic measures and social experiences (17). Participants read the information sheet, and completed the consent form and screening questions, via a Qualtrics survey, over email, or in person with a study researcher or clinical studies officer from participating NHS Trusts. Following informed consent, we ascertained eligibility via screening questions on current and previous diagnoses and self-identification, birth sex, and gender identity. We purposively sampled participants to include a mix of gender identities, ethnicities, and diagnosed versus self-identified, and suspected autism.

We recruited clinicians from NHS adult community mental health, forensic or private sector services, through the study team and local NHS Research and Development departments advertising the study by email, at clinical team meetings, at relevant conferences, and through our professional networks and snowballing from previous participants. We sent interested clinicians an information sheet, consent form, and a demographic questionnaire.

Sampling ended once we judged that data saturation and adequate information power had been achieved. Whilst our broad study aim reduced information power, our diverse sample with specificity for the studied issue, underpinning theoretical perspectives, and rich quality of researcher-participant dialogue increased information power (18).

### Qualitative interviews

JP and KB developed separate semi-structured topic guides for lived experience and clinician interviews, reviewed by our lived experience and clinician panel members (Online Supplementary Information). We asked lived experience participants how they came to know/why they think they may be autistic, whether they want to get an autism diagnosis and why, and whether they agree with their personality disorder diagnosis and why. We asked clinicians to share their experiences of differentiating autism and personality disorder in clinical practice. We asked all participants to discuss their experiences of facilitators and barriers to recognising autism in BPD-diagnosed people, including potential barriers identified from existing research (gender stereotypes, confusion between personality disorder and autism, communication difficulties, the limitations of current assessment methods), and any other factors identified by participants.

We audio-recorded the interviews and conducted them all via telephone or video conferencing, excepting one in-person lived experience interview. Interview duration ranged from 30 minutes to two hours (lived experience), or 20 to 60 minutes (clinicians).

### Thematic analysis

We analysed data inductively and reflexively (15), following Braun and Clarke’s (19) thematic analysis steps: data familiarisation, coding, theme generation and refinement. JP coded interview transcripts using NVivo 14 software (20). Coding and thematic development was reviewed and refined with KB.

### Ontology, Epistemology and Reflexivity

Through our critical realist stance, combining realist ontology with relativist epistemology (16), we considered how personal, social, and cultural context had influenced participants’ accounts and our interpretation. Lead interviewer and analyst JP has lived experience of personality disorder diagnosis and is a late diagnosed autistic individual. This potentially expedited rapport and understanding with autistic participants (9). Co-analyst KB has clinical research experience within personality disorder services. The authorship team embrace the neurodiversity paradigm in relation to autism (21). We appreciate critical stances on the personality disorder construct (22), whilst aiming to maintain a neutral and open stance on its validity in our work.

When interviewing, JP aimed to communicate a neutral stance on the constructs and processes involved in diagnosing autism and personality disorder, to encourage openness and transparency. JP used open, non-leading questions and neutral prompts to keep the narrative centred on participants’ own words. She made brief reflexive notes following each interview, and revisited them during coding, to distinguish participants’ language from assumptions rooted in her lived experience. During analysis, she actively sought divergent perspectives. She reviewed analysis regularly with KB and our clinician and lived experience panels, to ensure the analysis remained grounded in participants’ accounts.

## Results

Fifteen lived experience participants and fifteen clinicians participated. Figure 1 shows recruitment of lived experience participants.

We recruited eleven clinicians through NHS Trusts, three through our professional networks, and one via our clinician advisory panel. We contacted two further clinicians who had expressed interest, but they did not continue to interview.

Tables 1 and 2 summarise participants’ demographic and clinical/professional characteristics. Participants were from the North-West, North-East and South-East regions of England.

Formally diagnosed autistic participants were diagnosed between 0.5 and 14 years after their personality disorder diagnosis. Clinicians were psychiatrists, psychologists, nurse practitioners, service leads or peer support workers.

### Thematic analysis

We identified four main themes (Figure 2). Theme 1, ‘Borderline personality disorder: A sticky label based on spurious evidence’, addresses problems with the way diagnosis of BPD can be made and maintained by the system. Theme 2 ‘Seeing beyond BPD to autism’ outlines how apparent BPD symptoms can reflect undiagnosed autistic traits. Theme 3, ‘Overcoming barriers to autism assessment’, encapsulates difficulties in accessing autism diagnosis. Theme 4 details positive and negative ‘Consequences of recognising autism’. The themes are illustrated with quotes from lived experience (prefixed LX) and clinician (prefixed CL) participants.

### Theme 1: Borderline personality disorder: A sticky label based on spurious evidence

#### 1.1 : BPD: A diagnosis that does not fit

Almost all lived experience participants disagreed with, or were questioning, their BPD diagnosis. Over half spoke of their diagnosis being made at a young age, with minimal assessment and/or based on spurious evidence. Both clinicians and BPD-diagnosed people spoke about it being diagnosed at times of crisis, with self-harm or suicide attributed to BPD without consideration of alternative explanations.

“*I went into a unit after overdosing and I was just monitored, and I had a chat with a psychiatrist. And then they just came up with that, personality disorder. It was as quick and as simple as that*.” (LX302, disagrees with BPD, self-identified autism)

“*People aren’t rigorous enough… It’s just ‘So you’re behaving in a chaotic way, you’ve got a personality disorder’. Well, let’s think, have you always been like that? Were you managing? What happened? What were you like?*” (CL755).

Misdiagnosis was felt more likely when risk management was the focus.

“*A person may be presenting with high risk and so you see the risk behaviours and you’re trying to understand them, but you can’t see loads of other stuff that might lead to autism… And then, there’s a lack of curiosity when someone’s in a crisis, staff are worried, they’re trying to act and contain risk*.” (CL756).

Some participants also felt that bisexuality, tattoos, expressing dissatisfaction with mental health services, a history of trauma, or simply not fitting neatly into other diagnostic categories, contributed to BPD diagnosis. Additionally, questions asked as part of personality disorder assessment were experienced as unclear and vague, leading to participants answering yes without really understanding the question.

“*I remember him asking me if I feel empty - often empty inside, and I remember sort of like not really knowing what he meant, and I just said ‘Yes’*.” (LX300, disagrees with BPD, self-identified autism).

Participants felt clinicians were incentivised to diagnose BPD by service structures that necessitated this label in order to access psychological interventions such as dialectical behaviour therapy (DBT). Consequently, investigation of possible autism was deprioritised.

“*At the time it would have been considered a time of crisis coming off a suicide attempt…it was like ‘I do think that there are traits of Asperger’s in there… it’s not something we’re going to be able to explore….let’s get you into some DBT therapy, let’s sort the borderline…The Asperger’s…that’s not life threatening you’ll be okay, you seem to have managed so far.’*”(LX527, agrees with BPD, self-identified autistic).

Similarly, in forensic services, access to the support offered by the personality disorder pathway was contingent on difficulties being conceptualised under this diagnosis. These siloed service pathways could amplify a tendency to see the person through the lens of the service they were under, rather than holistically.

“…*We divide people up – we divide services up. People have to fit into the service rather than service fit around the person*.” (CL755)

For many, life transitions where the environment was problematic, or highly stressful life events, coincided with their BPD diagnosis. These included the menopause, infertility, serious illness in the family, or moving to university. BPD often did not feel like a fitting diagnosis once these stressors had been removed.

#### 1.2 A sticky label that cannot be questioned

A pre-existing BPD diagnosis frequently led to alternative explanations being overlooked. All participants shared examples of how potential autistic characteristics, such as rigid thinking, were interpreted as relating to personality disorder.

“*But then as I was under the personality disorder team it sort of moved to mixed personality disorder, including things like the obsessive compulsive with like the rigid thinking kind of and perfectionism, liking things a certain way…I feel they were very much looking at all my issues through that sort of personality disorder lens.*” (LX538, disagrees with BPD, waiting list for autism assessment).

Clinicians recognised the bias that came with this; how someone’s experiences can be interpreted as fitting within the label already ascribed, rather than considering alternative explanations.

“*I think because there’s that overlap, I think it’s understandable for clinicians to attribute certain sort of traits or presentations to that, personality disorder, because…that’s the diagnosis that’s already been given, rather than maybe digging into it a little bit more and finding out actually is this more about understanding what is the degree of emotional understanding, what is the degree of social understanding?*” (CL151).

Clinicians shared that some colleagues were reluctant to question historical diagnoses, and that patients questioning their BPD diagnosis were frequently seen as being in denial. Whilst one clinician agreed that patients’ rejection of their BPD diagnoses could be an external projection of their negative self-perception, most clinician interviewees disagreed with these perspectives but felt it was difficult to challenge colleagues.

“ ‘*She needs to accept it, she’s in denial’, and then it puts you in a really difficult position, because I don’t agree with that.*” (CL352).

Similarly, lived experience participants were very aware of clinicians’ negative perceptions around BPD and autism, consequently feeling unable to question their diagnosis or raise autism as a possibility.

“*‘Oh, here we go, it’s just another one of the EUPD girls, going, “Well, I actually think I’ve got this now”’…there’s a lot of stigma from a professional perspective there, which I’d be concerned about facing*.” (LX544, unsure if agrees with BPD, suspects is autistic).

Participants experienced their BPD diagnosis as removing their agency in determining what diagnosis best explains their experiences, negatively affecting their wellbeing.

“*I feel like I’ve literally got this diagnosis that I’m stuck with for the rest of my life now, that doesn’t seem true to me. And I think that probably actually made my mental health worse, because I was like, ‘But it doesn’t fit, I should be feeling this way, but I’m not. What’s wrong with me?’*” (LX304, disagrees with BPD, diagnosed autistic).

### Theme 2: Seeing beyond BPD to autism

#### 2.1 : Opening my eyes to autism

Many, but not all, lived experience participants described that learning about autism had been incumbent upon themselves. They learnt about it from television programmes or social media, or from diagnosed friends and family, followed by in-depth reading on the subject and completing screening questionnaires online. This led to the often surprising and initially jarring realisation that autism fitted closely with their experiences, sometimes much more clearly than BPD.

“*I started studying psychology …and when we were studying [autism], I kind of looking back realised that a lot of it felt more like applicable. And some of the things that I could kind of see being potentially related to a personality disorder were far more consistent with autism*.” (LX552, disagrees with BPD, suspects is autistic).

Conversely, a few participants spoke about clinicians having made the initial observation that they may be autistic. Again, this was often a surprising revelation.

“*I was in the local psychiatric hospital, and it was one of the staff there, that said, ‘Do you think you could be autistic?’ I was just like, ‘No, no, definitely not*…’ *even though I was in denial when she asked, I think just planting that seed and just, yeah, I think if someone would have done that during my first admission, it would have been very different*.” (LX304, disagrees with BPD, diagnosed autistic).

For clinicians, being open to considering undiagnosed autism came from confidence, personal and professional experience, and increased awareness. Several spoke of how this had changed over time for them.

“*I’m just trying to think of myself a few years ago…it wasn’t on my radar actually. I remember I could probably be judgmental and intolerant if people suggested other things, but I just completely missed if anybody was suggesting to me they might be autistic; I was so unaware I didn’t even hear it.*” (CL750).

Participants recommended joining up of autism and mental health services, and training about autism for those working in mental health services, as helpful for improving clinicians’ understanding and confidence in recognising autism.

#### 2.2 : Recognising autistic traits

Participants spoke about learning to recognise autistic traits, in themselves or in their patients. Developmental history was mentioned primarily by clinicians as being important in identifying autism. They spoke about recognising a history of autistic traits emerging in early childhood, including repetitive play, intense interests in certain toys or hobbies, systemising behaviours, and difficulties understanding relationships. Clinicians also spoke about the lifetime distressing effects of navigating a neurotypical social world with unrecognised autism.

All participants referred to sensory processing differences as a key indicator of possible autism. These included intense and aversive sensitivity to sounds, lighting, temperature, and textures such as clothing and food.

“*There are certain sensory experiences that come with being, let’s say, female, that make me extremely uncomfortable… And it’s possibly an explanation of why my eating disorder came at an onset of basically prepubescent age, because my body was starting to change*.” (LX562, disagrees with BPD, diagnosed autistic and ADHD).

Many participants discussed recognising autism-aligned differences in social interaction. These included a core difficulty in understanding neurotypical social communication, particularly implicit social cues and expectations, often combined with being overly trusting and easily taken advantage of. This was seen as uncharacteristic of BPD, which was instead associated by clinicians with hypersensitivity to social cues alongside a tendency to interpret others’ social behaviour as rejection.

“*She didn’t get the content of the conversation because…it was more implicit than explicit… being in a group you know and misunderstanding what is being said, or saying the wrong thing and being laughed at because of that, you know, those are things that not necessarily will happen in a personality disorder*.” (CL450).

Other autistic characteristics referenced included taking things literally, unusually direct communication, difficulty with large or noisy social situations due to sensory overload, a strong need for predictability and routine, highly focused interests, and difficulties in identifying and describing one’s emotions. Lived experience participants said clinicians often did not connect these to undiagnosed autism.

“*I ranted about my special interest for like 20-minutes. And my special interest was Pokémon. So, they’ve written my special interests like in my protective factors, but at no point in that have they gone like, maybe this is autism.*” (LX563, disagrees with BPD, diagnosed autistic).

Clinicians and lived experience participants also suggested that a history of multiple prior and changing mental health diagnoses, other neurodivergence such as dyspraxia or tics, and/or neurodivergent family members could be hallmarks of undiagnosed autism. Several clinicians suggested that difficulty benefiting from therapies for BPD, such as mentalization based therapy (MBT) or cognitive analytic therapy (CAT), could itself be indicative.

“*Say I’m doing CAT therapy and we’re trying to understand a pattern, a relational pattern or a behavioural pattern…Sometimes I’m wondering, is it something that person can’t change, rather than doesn’t want to change?…The capacity to understand or change something. It feels like a bit more kind of fixed*.” (CL351).

#### 2.3 : Looking beneath the surface

Both clinicians and lived experience participants highlighted that some characteristics and behaviours initially attributed to BPD could be explicable by undiagnosed autism. They felt that both diagnostic groups experience difficulties with emotional dysregulation and self-damaging coping behaviours, but that the precipitating factors underlying this are different. For instance, participants attributed emotional dysregulation in autistic people to unmet sensory needs, alexithymia, masking, fatigue, difficulties around change, and social differences.

“*I have mood swings but it’s more anxiety when things are thrust upon me, like being in a crowd, or changes, or when I can’t fix things…everything has to be in the right place. If somebody moves anything I get really panicky. So, I’ve compared the two, and they seem to be completely different things, that’s what sort of made me think am I autistic, do I have some traits of it?*” (LX302, disagrees with BPD, suspects is autistic).

Participants spoke about autism-aligned sensory processing differences being interpreted as BPD-aligned dissociation, with inferences made that difficulties concentrating were a reaction to emotional content rather than to sensory overwhelm.

“*They’d say, ‘Oh is that, because the conversation’s hard’ or whatever…and I think that they used to say like maybe that I was suffering from dissociation…And I would think, ‘No I just…’it [the clock] had like a dangly tail and it was a particularly loud and vivid clock that just used to always take my attention span away.*” (LX300, disagrees with BPD, self-identified autistic).

Conversely, clinician participants suggested that emotional distress may stem more from relational difficulties in people deemed to have BPD, linked to core beliefs following traumatic life experiences, and feelings of rejection and abandonment.

“*What I find in personality disorder…the trigger normally is some kind of social interaction…In people with autistic spectrum conditions [it] is not necessarily that clear. It might be more you know, an emotional state that they don’t know how to handle or express, you know, but not because of the interaction*.” (CL450).

Participants spoke about autism-aligned social differences being misinterpreted as symptoms of BPD, including misattribution as fear of abandonment.

“*I’ve had people tell me, or have clinicians tell me, that somebody is sensitive to abandonment and then, when you really listen it’s probably not that, it’s more that they find personal interactions very difficult and overwhelming and therefore, they have a reaction when that interaction ends*.” (CL750).

Similarly, they shared that autism-aligned difficulties in understanding neurotypical social communication, particularly implicit social cues and expectations, had sometimes been misattributed to BPD-aligned difficulties with mentalizing, or BPD-aligned unstable relationships.

“*Because I didn’t understand how relationships are meant to work…I did whatever they wanted and then obviously when I couldn’t cope with that, I had a bit of a meltdown.*” (LX500, unsure if agrees with BPD, diagnosed autistic).

Some participants also spoke about an autistic tendency to intensely focus on a few relationships being misunderstood as BPD-aligned idealisation. Similarly, use of alcohol or substances, or risky sexual behaviour had been attributed to BPD-aligned impulsivity, when it could actually be an autistic person using substances to manage social situations, or being vulnerable to exploitation and easily led to do what others wanted. Autism-aligned focused interests, including psychology, fashion, and women’s rights, could also be misinterpreted when viewed through the lens of personality disorder. In one example, a highly focused interest in feminism was misinterpreted as BPD-aligned anger towards males.

People identifying with both diagnoses found this very confusing, finding it difficult to pick apart which aspects of themselves were explained by which diagnosis. Participants felt co-occurring ADHD could further contribute to diagnostic confusion, due to potentially counteracting autistic traits.

“*The autism traits of, you know, being extremely organised, needing structure, needing routine, almost counteract the impulsivity, the chaotic-ness of the ADHD…there are almost like ‘key features of autism’ that I don’t demonstrate in a way because my ADHD essentially counteracts them.*” (LX562, disagrees with BPD, diagnosed autistic and ADHD).

Another complicating factor raised by participants was whether a lifetime of masking autistic traits, coping with unmet autistic needs, social exclusion, and bullying (‘neurodivergent trauma’), could lead autistic people to develop BPD-aligned characteristics such as emotional dysregulation and rejection sensitivity. Participants wondered whether this was made even more likely when an autistic person had experienced sexual or physical violence. Some felt, therefore, that autism and BPD could often coincide. Others disagreed with the BPD construct and felt it was more helpful to formulate experiences as a combination of autism alongside complex trauma.

“*I think the difficulty both clinicians and clients have is that is that little word ‘and’. You know that there could be neurodiversity issues which makes you particularly sensitive to these traumas, and you’ve been traumatized a lot as well. And it might be both*.” (CL751).

### Theme 3: Overcoming barriers to autism assessment

#### 3.1 Seeing through stereotypes about autism

Both participant groups expressed that lack of knowledge about different autism presentations could be a barrier to recognition. Participants described some clinicians as lacking awareness of how autism can present in women and AFAB people, and/or autistic people with a more ‘internalised’ presentation. They spoke about autistic individuals learning to ‘mask’ or disguise their autistic traits to fit in, making autism assessments difficult. To overcome this, clinicians spoke about moving away from observational autism assessments, such as the ADOS, and using more self-report questionnaires alongside people’s accounts of their experiences. They also spoke about being alert to signs of compensatory behaviour disguising autistic traits, spending more time with someone as patients could struggle to mask their autistic traits for extended periods, and doing assessments in pairs to facilitate observation of more subtle autistic behaviours.

“*We have diagnosed her. She doesn’t score in the ADOS…I did a very much longer assessment than normal…after one hour and 40 minutes…her masking skills started to drop.*” (CL450).

Clinicians and lived experience participants experienced stereotypes of autism as characterised by intellectual difficulties. They felt it was important to be aware that autistic people could excel in some areas of life and really struggle in others. In areas where a person was perceived to be functioning well, they cautioned it was important to assess the effort and costs involved for the person, such as attendance at social events being followed by exhaustion and shutdown.

“*It’s not just about, ‘Yes, I can do this or no, I can’t do things’…It’s the cost of being able to do something, isn’t it? And the impact that cost might have on other areas of life; and that takes a bit of time to unpick.*” (CL151).

#### 3.2 Overcoming a lengthy and complicated diagnostic process

Lived experience participants were very aware of the long waiting lists for an autism assessment and/or the cost of seeking private assessment. For some, this put them off seeking an official diagnosis. This was particularly the case for participants who, jaded by previous negative experiences with services, did not want to put themselves through the anticipated difficult process of persuading clinicians to refer them for an assessment. They feared they would not be taken seriously and worried about the damage to their mental health that this would cause.

“*I didn’t really trust my clinical team to take me seriously because of the way they responded to me when I had asked questions about other things in the past.*” (LX539, unsure if agrees with BPD, diagnosed autistic).

Both groups shared that the need to involve someone who knew the person well as a child could be a major barrier, especially where parents were elderly or deceased. Further, some participants did not trust their parents to support the diagnostic process, did not feel their parents knew them well enough to give an accurate account, or did not feel comfortable being in contact. Not being able to get a developmental history had prevented some participants from being diagnosed as autistic. Clinicians held mixed views on the necessity of this, with some saying it was essential for diagnosis. Others said they could instead use personal or sibling accounts of childhood, alongside reports from people who know them well in adulthood, with team consensus used to make decisions in cases where information was missing or unclear.

### Theme 4: Consequences of recognising autism

#### 4.1 I know who I am and what I need

Diagnosed or self-diagnosed autistic participants were unanimous that recognising autism had led to a more positive and clear self-understanding, as well as improvements in psychological well-being and quality of life.

“*…the BPD diagnosis it felt very much like I was being told, ‘Okay you’re like this because you’re not trying hard enough’, so that just meant that I pushed myself to try harder and harder and harder and led myself to burning out over and over and over again. Whereas with, now I have the autism diagnosis, I can say actually, no, that environment is not compatible with my needs*.” (LX539, unsure if agrees with BPD, diagnosed autistic).

This also facilitated improved relationships with others, and finding different ways of responding to areas of life that had previously been misunderstood, such as understanding sensory needs underlying disordered eating. Recognition of autism allowed for better decisions around what psychological therapies may be appropriate, as well as where interventions involving groups could be more difficult. Clinicians shared concerns that un-adapted therapies could be harmful, for instance by not recognising autistic individuals’ different ways of mentalizing.

“*They told her that they didn’t think that MBT was going to help her because of her difficulties with reading social cues and some of her literal interpretation of some of the meaning of MBT*.” (CL353).

### 4.2 A two-way diagnosis of exclusion

A minority of participants, however, raised concerns that an autism diagnosis could inhibit access to psychological interventions, particularly with a co-occurring BPD diagnosis. Others raised concerns about stigma associated with autism.

“*Since I got both diagnoses, I’ve not had as much contact with clinicians because it’s made me ineligible for most mental health help…because I’m now officially a complex case.*” (LX525, unsure if agrees with BPD, diagnosed autistic).

“*If we diagnose them with autism, then nobody else wants to take them…and the fact that they have a personality disorder and autism excludes them from services.*” (CL751).

## Discussion

This study is the first to integrate clinician and lived experience perspectives on barriers and facilitators to recognising autism in BPD-diagnosed people. We identified four themes, spanning the barriers people face in questioning their BPD diagnosis (Theme 1), the process of recognising and distinguishing autistic traits from BPD (Theme 2), overcoming barriers in the autism assessment process (Theme 3), and the consequences of recognising autism (Theme 4).

### Diagnosis and questioning of BPD

Our findings suggest that a BPD diagnosis can overshadow identification of autism, particularly in women/AFAB people. Relatedly, we know that autistic females are more likely to have other mental health diagnoses compared to males, and that personality disorder is one of the most frequent prior diagnoses given to late-identified autistic people (1). Our participants felt that overshadowing is particularly likely when BPD is diagnosed at a young age or during times of crisis. The potential merits and harms of diagnosing BPD in adolescents has been hotly debated (25). Periods of life transition, stress and/or significant hormonal shifts such as menopause, are known to be very stressful for autistic people and to lead to mental health crises (26) – potentially a high-risk time for misdiagnosis with BPD.

Our interviewees spoke to the systemic factors reinforcing diagnosis of BPD over autism, including the BPD diagnosis as a gateway for access to psychological therapies such as DBT and MBT (27). This adds nuance to the perspective shared by some autistic people that services are motivated to diagnose BPD over autism based on ‘cost-effectiveness’ (12), instead highlighting the role of more beneficent clinical intentions. Aligning with previous research, participants shared the difficulties they experienced when challenging a pre-existing personality disorder diagnosis – at times even having their questioning pathologized as an aspect of personality disorder (12, 13).

### Differentiation of autism and BPD

Our findings reinforce that rapid diagnosis of BPD based solely on emotional dysregulation, self-harm and suicide attempts, without the realisation that these could potentially be attributed to undiagnosed autism, can be problematic (11, 12). In line with previous thinking and research, our study highlights that emotional overwhelm linked to sensory overload or loss of routine can be misattributed to BPD, alongside autistic experiences of shutdown in response to sensory overload being misinterpreted as BPD-aligned dissociation, and autistic social differences being misinterpreted as BPD-aligned mentalizing difficulties, relationship instability and fear of abandonment (3, 12, 28).

### Identification and diagnosis of autism

Many of the barriers we identified are consistent with the wider literature on factors that prevent recognition and diagnosis of autism, including lack of autism awareness and dismissive attitudes amongst clinicians (13), alongside lengthy waiting lists for autism assessments (29). Our findings align with the well-known difficulties in identifying autism in women and AFAB people (30, 31). We also highlight the challenges around obtaining a developmental history from a childhood informant, which can be unreliable or impossible for older adults or for people with a disrupted and traumatic childhood (32).

### Implications for clinical practice and further research

Our study points to the importance of recognising autism in people with a BPD diagnosis, with our participants unanimously finding this brought a more positive and clear self-understanding, alongside improved well-being. Participants’ accounts reinforce that BPD diagnoses should not be made without full consideration of differential diagnoses such as autism. Clinicians should learn how to recognise autism – particularly more internalised presentations that can be characteristic of autistic women/AFAB people. By better understanding the similarities and differences between autism and BPD, and having autism ‘on their radar’, identification of autism can be improved, and misdiagnosis reduced. Our findings highlight the value of co-working between autism and personality disorder services, with each offering alternative perspectives and questioning siloed thinking. We suggest the importance of trusting individuals as knowers of their own experiences (‘epistemic justice’), thus taking them seriously when they question whether they may be autistic (22).

Beyond BPD, wider problems with the autism assessment process must be addressed. Future research should develop and test revised autism diagnostic processes that better reflect internalised or female autism presentations. UK guidance on the necessity of childhood informants is currently mixed (33, 34). The validity of alternative ways of gaining developmental histories for adults requires investigation, so that individuals for whom this is not possible are not excluded from diagnosis.

Once it is recognised someone with a BPD diagnosis is autistic, a question remains as to whether the BPD diagnosis was incorrect. A growing body of lived experience and clinical work argues BPD is never a valid diagnosis (7, 22). Many of our participants also felt uncertain about their BPD diagnosis, yet felt unable to question it, or were pathologised for doing so. We argue that questioning one’s diagnosis should be taken seriously and should not be pathologised as symptomatic of BPD, which is likely to further iatrogenesis for this patient group.

Concerningly, our findings suggest that co-occurring autism and BPD may exclude individuals from evidence-based psychological therapies, and that clinicians may avoid or deprioritise diagnosing autism in fear of this. Given that a BPD diagnosis is experienced as harmful by some recipients (22), we believe that receipt of psychological therapies such as DBT and MBT should not be contingent on this. Further, allowing transdiagnostic access would enable these therapies to benefit other diagnostic groups also known to experience emotional dysregulation and mentalizing difficulties, such as autistic and ADHD individuals (35, 36, 37). However, our participants shared that specialist therapies for BPD were not always helpful for autistic people, sometimes reinforcing a negative self-perception, pushing them to change themselves rather than make needed adaptations to their environment, or failing to realise their inherent mentalizing differences. Relatedly, there is growing recognition that psychological therapies which do not consider autistic needs, can cause iatrogenic harm, and a loss of trust in services may ensue (10). There is an urgent need to adapt and test evidence-based interventions to better meet the needs of autistic individuals.

### Limitations and Critical Realism

Whilst our sample reflects the gender diversity of autistic people, 80% were white British and the sample were highly educated, reducing transferability to under-represented groups. Participation in the research, participants’ accounts within interviews, and our interpretation of these, were likely shaped by current societal discourses that are sceptical of BPD diagnoses and that embrace the current rise in autism diagnosis (38), in contrast to more sceptical viewpoints (39). Participants’ accounts within interviews may have been further shaped towards neuroaffirming and BPD-sceptical viewpoints, through disclosure of the interviewer’s status as a late diagnosed autistic individual with a co-occurring BPD diagnosis. As detailed in the Reflexivity section, the open and neutrally worded interview questions, and our reflexive team-based analysis alongside presentation of divergent perspectives, aimed to mitigate these influences.

## Conclusion

Combining clinical and lived experience accounts, we identified that recognising autism in someone with a BPD diagnosis can be both challenging and life-changing. Improving clinician awareness of different presentations of autism and of differential diagnosis from BPD, is likely to reduce misdiagnosis. Being open to questioning pre-existing diagnoses, and improving transdiagnostic access and neurodiversity affirming adaptations to evidence-based psychological therapies, is also vital.

## Acknowledgements

We would like to thank all of our participants for their time and expertise, and we are grateful for the advice and feedback received from our lived experience advisory panel (Jenn Layton Annable, Jo Barnes, Eva Broeckelmann, Eloise Curtis, Peg Digitalis, Sarah Labovitch, Anonymous) and our clinician advisory panel (Mike Crawford, Tennyson Lee, Silvia Murguia, Rachael Line, Richard Smith, Richard Pender, Harriet Fletcher, Simon Graham).

## Author contributions

Conceptualisation, Funding acquisition, Methodology, Writing – review & editing: KB, ET, ES, WM, RM, JP. Investigation: JP, KB, ET. Data curation, Project administration, Formal analysis, Visualisation: JP. Writing – original draft: JP, KB. Supervision: KB.

## Statements and declarations

### Ethical considerations

This study was approved by the West Midlands - South Birmingham National Health Service (NHS) Research Ethics Committee on 14th January 2022, (REC ref. 21/WM/0287, IRAS 307912). All participants provided written or electronic informed consent prior to enrolment in the study. This research was conducted ethically in accordance with the World Medical Association Declaration of Helsinki.

### Consent to participate

All participants provided written or electronic informed consent prior to enrolment in the study.

### Data availability

The datasets generated during the current study are not publicly available due to privacy concerns but are available from the corresponding author on request.

